# Guidance for the Reporting of Bibliometric Analyses: A Scoping Review

**DOI:** 10.1101/2024.08.26.24312538

**Authors:** Jeremy Y. Ng, Henry Liu, Mehvish Masood, Niveen Syed, Dimity Stephen, Ana Patricia Ayala, Michel Sabé, Marco Solmi, Ludo Waltman, Stefanie Haustein, David Moher

## Abstract

Despite the growth in the number of bibliometric analyses published in the peer-reviewed literature, few articles provide guidance on methods and reporting to ensure reliability, robustness, and reproducibility. Consequently, the quality of reporting in existing bibliometric studies varies greatly. In response, we are developing a preliminary **Guidance List for the repOrting of Bibliometric AnaLyses (GLOBAL)**, a reporting guideline for bibliometric analyses. This paper outlines a scoping review that aims to identify and categorise bibliometric recommendations from the literature to develop an initial list of candidate items for the GLOBAL. Five bibliographic databases, three preprint servers, and grey literature were systematically searched. Twenty-three out of 48,750 records fulfilled the inclusion criteria. Six documents contained bibliometric reporting recommendations based on a complete or partial literature review; all other sources (n = 17) contained opinion-based recommendations. A 32-item recommendation list that will inform the development of the GLOBAL was created. A paucity of evidence-based studies on bibliometric reporting exists in the literature, supporting the need to create a reporting guideline for bibliometric analyses. The next step in the GLOBAL project will focus on conducting a two-round Delphi study to achieve consensus on which of the 32 items should be included in GLOBAL.

## 1. Background

A bibliometric analysis is a method of evaluating and analysing the characteristics of a particular research area or field, or the research output of an individual, a group, or an institution, by examining and quantifying various aspects of their published works, such as the number of publications and citations, or collaboration patterns (De Bellis, 2014).

Bibliometric studies are typically performed using specialised databases and tools that allow researchers to search, retrieve, and analyse bibliographic data, such as article and journal titles, abstracts, author names, publication dates, as well as cited references and citing documents of journal articles, conference papers, and other published works (AlRyalat et al., 2019; Donthu et al., 2021; Sugimoto & Larivière, 2018). Bibliometric analyses can provide valuable insights into research trends and emerging topics, knowledge flows, and collaboration patterns. They can also identify key players such as researchers, institutions, countries, or journals and make visible the scope and impact of the work of these players (Donthu et al., 2021; Linnenluecke et al., 2020).

Bibliometric analyses can be conducted at various levels of granularity, such as by a single researcher, a department/institution, or an entire field or discipline. The specific metrics and indicators used in a bibliometric analysis will depend on the particular research question and the type of data being analysed (AlRyalat et al., 2019; Donthu et al., 2021; Sugimoto & Larivière, 2018). Most bibliometric analyses are based on two basic units: publications (e.g., journal articles, conference proceedings papers, book chapters, preprints) to represent scholarly outputs, and citations (i.e., formal references from one publication to another) to reflect knowledge flows between them.

Bibliometric methods have become an increasingly important tool for research evaluation, particularly in the context of grant proposals, promotions, institutional assessments, and other types of academic evaluation (Donthu et al., 2021; Ellegaard & Wallin, 2015; Koo & Lin, 2023). While some policymakers and evaluators are trying to reduce the role of bibliometric methods in research evaluation (CoARA, n.d.), bibliometric analyses continue to grow within various specialized fields such as library and information sciences, business, and health sciences (Donthu et al., 2021; Ellegaard & Wallin, 2015; Koo & Lin, 2023).

However, despite this increase in popularity, there is currently a lack of evidence-based guidance on how to report a bibliometric analysis (Cabezas-Clavijo et al., 2023). Establishing standard reporting guidelines for these types of studies is crucial for improving their quality and validity. Such guidelines can positively influence how researchers plan and execute their work (Moher et al., 2010), thereby ensuring that published work is complete and transparent (Gagnier et al., 2013). As a first step to address this gap, we are developing a reporting guideline for bibliometric analyses, known as the **Guidance List for the repOrting of Bibliometric AnaLyses (GLOBAL)**. A reporting guideline can take the form of “a checklist, flow diagram or explicit text to guide authors in reporting a specific type of research, developed using explicit methodology” (Moher et al., 2010, p. 1). Our aim in creating the GLOBAL is to enable increased transparency, more complete and thorough reporting, and ultimately more rigorous bibliometric analyses.

As described in our protocol (Ng et al., 2023), the development of the GLOBAL project was informed by the “How to develop a reporting guideline” toolkit, provided by the EQUATOR Network (EQUATOR, 2018c), and guidance for developing reporting guidelines created by Moher et al. (2010). When creating a reporting guideline, a review of the literature must be conducted to identify previous relevant guidance, seek relevant evidence on the quality of reporting in published research articles, and identify key information related to potential sources of bias in relevant studies (EQUATOR, 2022; Moher et al., 2010; Ng et al., 2023). This scoping literature review may additionally serve to generate a candidate list of items for the reporting guideline, such as in the case of the ACcurate COnsensus Reporting Document (ACCORD; Zuuren et al., 2022). The preliminary list of candidate items would then undergo refinement, including, but not limited to, a Delphi exercise and face-to-face consensus meeting (Moher et al., 2010). The GLOBAL was developed in accordance with the aforementioned steps (EQUATOR, 2022; Ng et al., 2023): a scoping review (the present analysis) was conducted to identify relevant reporting guidance for bibliometric analyses and generate a preliminary list of candidate items, followed by the administration of a Delphi exercise (to be described in a subsequent report) to further develop this list. Consequently, the present study is a scoping review of peer-reviewed literature, grey literature, and preprint servers that aims to identify and categorise bibliometric reporting recommendations with the ultimate goal of developing an initial list of candidate items for the GLOBAL reporting guideline. This scoping review therefore focuses on asking the following question: What recommendations exist for reporting bibliometric analyses?

## 2. Methods

### 2.1. Approach

This scoping review gathered reporting recommendations for bibliometric analyses from peer-reviewed literature, grey literature, and preprint servers to develop an initial list of candidate items for the GLOBAL reporting guideline. This analysis occurred in consultation with a steering group of bibliometricians (LW, MSabé, MSolmi, and SH) and reporting guideline experts (DM). This scoping review was conducted in accordance with the methods outlined in the Joanna Briggs Institute (JBI) Manual for Evidence Synthesis (Peters et al., 2020), which describes a multistep process of search strategy development, evidence source screening and selection, data extraction, and data analysis and presentation.

### 2.2. Transparency Statement

The GLOBAL project was registered on the EQUATOR library of reporting guidelines (EQUATOR, 2022). The final protocol, study materials, and data were registered on the Open Science Framework (OSF, n.d.) in Ng et al. (2024). We followed the Preferred Reporting Items for Systematic Reviews and Meta-Analyses extension for Scoping Reviews (PRISMA-ScR; Tricco et al., 2018) in reporting our methods and findings.

### 2.3. Search Strategy

#### Bibliographic Database Searches

An information specialist (APA) searched the following databases from their dates of inception to July 28, 2023: MEDLINE and MEDLINE in Process via OVID, EMBASE Classic + EMBASE via OVID, APA PsycINFO via OVID, Web of Science Core Collection, and Scopus. A search strategy was developed in OVID MEDLINE using appropriate controlled vocabulary by one member of the research team (JYN) and was further refined with input from APA (see Appendix A for search strategy). The OVID MEDLINE search strategy was peer reviewed using the Peer Review of Electronic Search Strategies (PRESS) tool (McGowan et al., 2016) by an external information specialist. The peer-reviewed OVID MEDLINE search strategy was then adapted as required for application in the other databases (**Appendix A**). There were no restrictions by publication type or date. All searches were limited to documents published in English. The searches used a multi-string approach, combining the concepts of bibliometrics and reporting. All searches were retrieved on July 28, 2023.

#### Preprint Servers and Grey Literature Search

We conducted a search of preprint servers and grey literature (e.g., blogs released by researchers, websites of bibliometrics-related organisations and associations) to capture guidance documents that were not published as traditional academic articles. Sources of eligible grey literature were initially sourced by JYN, LW, and SH and then reviewed by the other authors (DS, MSabé, and MSolmi). The complete list of grey literature sources can be found in **Appendix B**.

Each website (see **Appendix B** for website links) was searched on February 20, 2024, using the following search term: “guidance on reporting bibliometric analyses.” The search bar was used if it was present on a given website. If no search function was present, the site was searched via Google (e.g., “site: website.com” + “guidance on reporting bibliometric analyses”). Separate from these websites, general Google searches were conducted on February 21, 2024, using the aforementioned search term (i.e., “guidance on reporting bibliometric analyses”). Searching Google facilitated the identification of relevant grey literature sources outside of our generated list of organisations. The first 100 search results were reviewed for eligibility. This threshold was selected as it captured the most relevant sources while maintaining a manageable number of records to screen.

Finally, searches were conducted on three preprint servers commonly used by members of the bibliometrics community: 1) arXiv (ArXiv, n.d.); 2) OSF Preprints (OSF Preprints, n.d.), which hosts the infrastructure for multiple preprint servers; and 3) Zenodo (Zenodo, n.d.).

The same search term (i.e., “guidance on reporting bibliometric analyses”) was used, and the first 100 results from each repository were reviewed for eligibility. The Zenodo search was completed on February 20, 2024, while the OSF Preprints and arXiv searches were conducted on February 23, 2024, and March 14, 2024, respectively.

### 2.4. Eligibility Criteria

The eligibility of the sources was based on a PCC (participants, concept, context) framework (see **Table 1** for screening criteria).

**Table 1:** Screening Criteria.

| Section | Criteria |
| --- | --- |
| **Aims and Objectives** | To conduct a scoping review of recommendations that exist for the reporting of a bibliometric analysis, which will inform the development of a reporting guideline for bibliometric analyses. |
| **Screening Criteria** | *Participants*<br>This criterion refers to study subjects or groups of interest. This review had <u>no restrictions based on participant characteristics</u>. The criterion of participants is likely not applicable to many literature sources that will be searched in this review.<br>*Concept*<br>This criterion refers to the core idea that was examined by this scoping review. This review focused on <u>recommendations for the reporting of a bibliometric analysis</u>. For this review, recommendations could be found in a range of eligible academic and grey literature sources (e.g., primary research, reviews, professional newsletters). Recommendations of interest took many different forms, oftentimes without explicit mentions of the terms “recommendation” or “reporting”.<br>*Context*<br>This criterion refers to the surrounding circumstances or setting of a given scoping review. This review had <u>no restrictions based on context</u>, such as cultural, geographic, or sociodemographic factors. The criterion of context was not applicable to many literature sources that will be searched in this review. |
| **Sources of Evidence Selection** | *Inclusion Criteria*<br>• Academic literature (e.g., primary studies, reviews, meta-analyses, commentaries) was included.<br>• Preprints were included.<br>• Grey literature sources (e.g., blogs, websites, newsletters) were included.<br>• Sources must be in the English language.<br>*Exclusion Criteria*<br>• Bibliometric analyses were excluded.<br>◦ While it is conceivable for a bibliometric analysis article’s author to comment on the current lack of reporting guidelines, the large number of bibliometric analyses that the search strategy returned rendered the full text review of every bibliometric analysis infeasible.<br>◦ An exception, however, is that if the bibliometric analysis’ title or abstract made specific mention of making reporting recommendations, it was included for full text review.<br>• Conference abstracts were excluded.<br>• Study protocols were excluded. |
| **Title/Abstract Screening Questions** | 1. Does the title or abstract mention recommendations (e.g., advice, suggestions, guidance, best practices, proposals, etc.) for the reporting of a bibliometric analysis?<br>2. Is the source a bibliometric analysis, conference abstract, or study protocol?<br>3. Is the source in English? |

### 2.5. Study/Source of Evidence Selection

#### Bibliographic Database Evidence Selection

All references from database search results were entered into an Endnote file (EndNote, 2013) for processing, and then uploaded to DistillerSR (DistillerSR, n.d.) for deduplication and screening. Endnote (EndNote, 2013) is a reference management software that can export references to DistillerSR (DistillerSR, n.d.), which is an online screening and data extraction workflow tool for conducting reviews.

Two reviewers (HL and JYN) conducted the title/abstract screening of 10% of the retrieved records, independently and in duplicate. During screening, records were sorted by DistillerSR Artificial Intelligence SYstem (DAISY) re-ranking. The DAISY feature uses natural language processing and the reviewers’ previous screening patterns to continuously predict which screened records are potentially the most relevant, before ranking unscreened records in order of relevance (DistillerSR, n.d.; Hamel et al., 2020). Following manual screening (HL and JYN), the title/abstract screening of the remaining 90% of the retrieved records was completed through DistillerSR’s AI Screening tool (Hamel et al., 2020). We opted to use the

DistillerSR AI Screening tool due to the large number of records (>45,000) retrieved from the database search. This decision was informed by previous literature, where a review conducted by Burns et al. (2021) to test the accuracy of AI screening tools determined that DistillerSR’s AI Screening tool has a sensitivity of 1.0 and a specificity ranging from 0.9 to 1.0.

DistillerSR’s AI Screening tool provides a score from 0 to 1 for each screened record to predict the likelihood that the record is relevant based on the previous human-based screening choices. A score of 0 indicates that a record can be excluded with very high confidence, while a score of 1 indicates that records can be included with very high confidence. It was determined *a priori* that documents eligible for inclusion would require a score greater than 0.5.

Two reviewers (HL, DS) then conducted full-text screening independently and in duplicate. All screening conflicts were resolved by consensus or, when necessary, third-party arbitration (JYN).

#### Preprint and Grey Literature Evidence Selection

The preprints and grey literature were assessed for inclusion via title/abstract screening by two reviewers (MM and NS), independently and in duplicate, after training by previous screeners (JYN and HL) to ensure consistency. Full-text screening of these documents was then similarly conducted by two reviewers (MM and NS), independently and in duplicate. Preprints and grey literature search results were deduplicated. All screening conflicts were resolved by consensus or, when necessary, third-party arbitration.

### 2.6. Data Extraction

#### Data Extraction Form Development and Pilot Testing

Prior to full data extraction, a preliminary data extraction form was developed by HL, JYN, MM, and NS, before being reviewed and modified by DM, LW, and SH. Two reviewers (MM and NS) participated in the pilot testing of the data extraction form. As part of the pilot exercise, each reviewer independently extracted data from a sample of five eligible full texts, including bibliographic, preprint, and grey literature sources. Once completed, the authors met to compare results, discuss and resolve conflicts, and revise the data extraction form as needed. This revised form was then used for all remaining extractions.

#### Full Data Extraction Phase

For each eligible source, the following information was extracted (as applicable): title; authors; digital object identifier (DOI) or uniform resource locator (URL); year of publication; country of first author; document objective; study design or source type; journal subject scope; whether a source gave special focus to non-bibliometric subjects; whether a document gave special focus to bibliometrics for use in hiring, promotion, or tenure; whether a source had bibliometric analysis reporting recommendations; whether reporting recommendations were evidence-based (i.e., if sources adhered to EQUATOR’s “How to develop a reporting guideline” toolkit [EQUATOR, 2018c]); manuscript sections to which reporting recommendations apply; sources informing the development of reporting recommendations; whether a document provides reporting examples; and conceptual frameworks suggested to understand bibliometric analyses. Crucially, all recommendations provided from each eligible source pertaining to the reporting of a bibliometric analysis were extracted.

Two reviewers (MM and NS) conducted the data extraction of all eligible full texts independently and in duplicate. Once data extractions were completed, reviewers met to compare their findings, discussing and resolving any discrepancies via consensus or, when necessary, third-party arbitration (JYN).

#### Collection of Items and Synthesis

Extracted bibliometric reporting recommendations from documents that met the inclusion criteria were categorised by HL, MM, and NS. Similar recommendations were combined based on categories to create candidate items for the GLOBAL reporting guideline (JYN, HL, MM, and NS). Iterative discussions with the research team resulted in consensus on the item’s inclusion, the section to which an item belonged (i.e., if the recommendation pertains to the ‘title’, ‘abstract’, ‘introduction’, ‘methods’, ‘results’, ‘discussion’, or ‘other’ section of the reporting guideline), and the phrasing of candidate items for the GLOBAL. Bibliometrics specialists on the steering committee (DS, LW, SH, MSabé, MSolmi) were also provided an opportunity to include items that were not directly addressed by the included studies but were perceived as necessary to enhance the rigour of bibliometric reporting.

### 2.7. Data Analysis and Presentation

The results of the search strategy and the study inclusion processes are fully reported and presented in a PRISMA flow diagram. Included documents are summarised quantitatively through frequencies of extracted item characteristics (e.g., year of publication) and presented in narrative and tabular form. Similarly, a list of all generated bibliometric reporting items for the preliminary GLOBAL checklist is presented narratively and tabularly.

## 3. Results

### 3.1. Screening and Search Results

A total of 48,750 documents were identified from all sources (48,315 from databases, 400 from preprint servers, and 35 from grey literature sources). Following the removal of duplicates (n = 2,904), a total of 45,411 records remained. Members of the research team screened 4,395 out of 45,411 of these remaining records, while DistillerSR’s AI screening tool was used to screen the remainder (n = 40,780). A total of 146 documents from the human screening process and one document from the automated screening process remained following this step. It was anticipated that limited records would be identified during automated screening due to the use of the DAISY re-ranking system, which resulted in most of the ’relevant’ sources being addressed during human screening, leaving the less ’relevant’ sources for automated screening. All documents were retrieved (n = 147) and were screened for eligibility. A total of 13 documents from the database search met the eligibility criteria and were included in the present review. In contrast, following the screening of retrieved preprint and grey literature records (n = 435), a total of 10 preprint server and grey literature records met the inclusion criteria and were included in the present review. See **Figure 1** to view the PRISMA flowchart diagram. Refer to Ng et al. (2024) to see title-abstract and full-text screening decisions, as applicable, for articles retrieved from bibliographic databases and grey literature.

**Figure 1:**
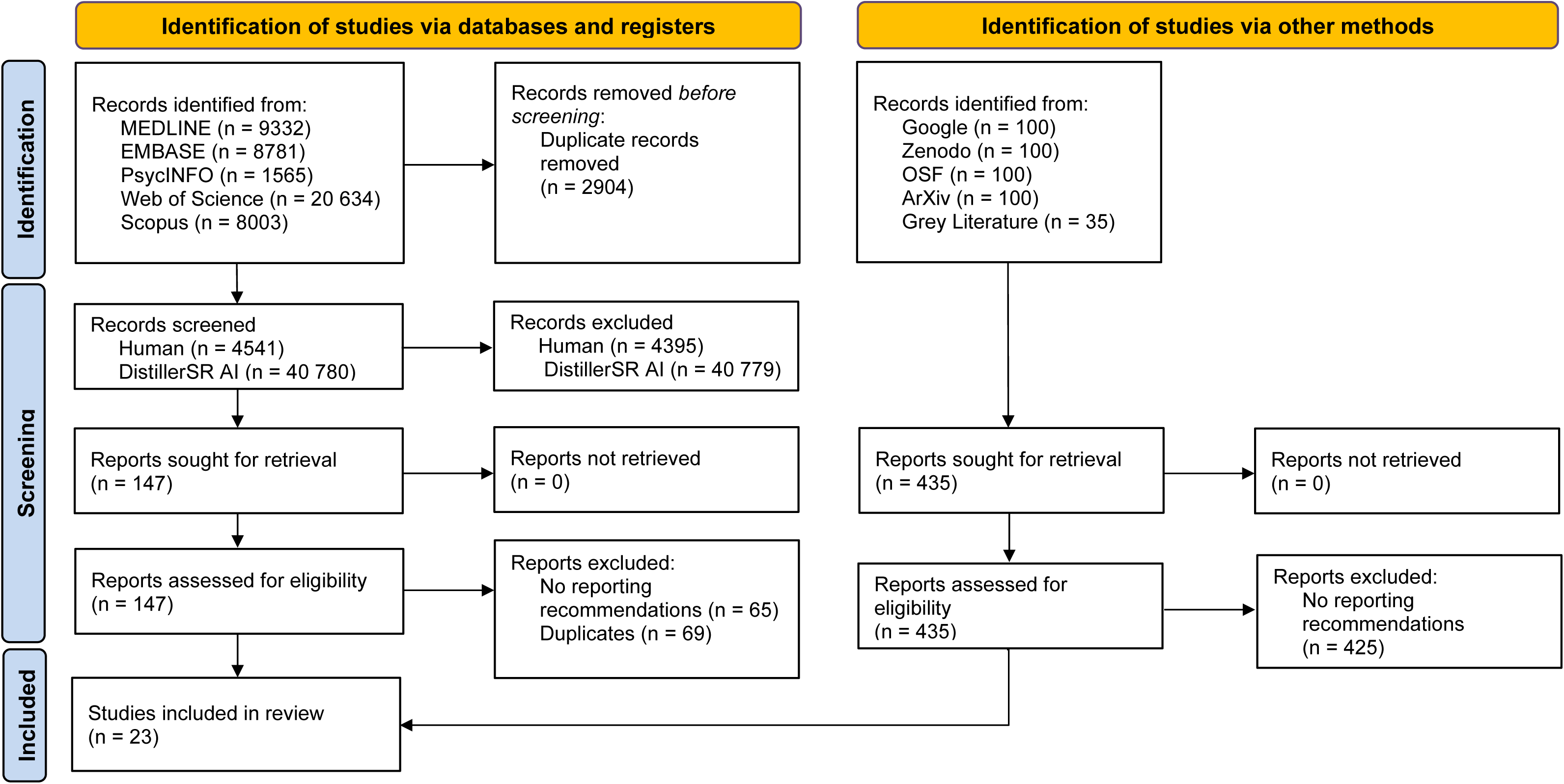
Preferred Reporting Items for Systematic Reviews and Meta-Analyses (PRISMA) Diagram

### 3.2. Characteristics of Included Studies

Following screening, a total of 23 documents met the inclusion criteria, 13 of which were from databases and 10 from preprint servers and grey literature sources. The study characteristics of the included documents are summarised in **Table 2**. A variety of study designs and/or source types were present within the included records. While the majority were guidance papers (n = 10), there were also websites (n = 2), reporting guidelines (n = 2), conference guidance papers (n = 2), a book chapter (n = 1), an editorial (n = 1), a LinkedIn article (n = 1), an opinion piece (n = 1), a research paper (n = 1), a university declaration (n = 1), and a white paper (n = 1). All except one source provided a year of publication; these documents were published between 1994 and 2023. The one source that did not provide a year of publication was a website that was last updated in 2024.

**Table 2:** Characteristics of Included Studies.

| Author and Year | Country of first author | Objective/Aim (as described by the document) | Study Design/ Source Type | Journal Subject Scope | Did the document give special focus to any non-bibliometric subject matter? | Did the document give special focus to the use of bibliometrics for hiring, promotion, or tenure of researchers for institutions? | Are reporting recommendations evidence-based<sup>a</sup>? | Does the study conduct an example bibliometric analysis to provide reporting examples? | Number of distinctive Bibliometric Reporting Recommendations | Did the document describe a conceptual framework of understanding different types of bibliometric analyses? |
| --- | --- | --- | --- | --- | --- | --- | --- | --- | --- | --- |
| Bornmann et al. (2014) | Germany | The aim of this chapter is to describe standards for applying bibliometric methods in the evaluation of research institutes in the natural sciences. | Book Chapter | Natural Sciences | No | No | No | Yes | 4 | No |
| Boyack et al. (2022) | USA | We provide suggestions as to how the bibliometric community might improve the state of the art in a way that can be used by non-experts. | Conference Guidance Paper | N/A | No | No | Partially | No | 8 | No |
| Byl et al.<br>(2016) | Canada | Not explicitly provided by source.<br>(To suggest bibliometrics recommendations to researchers, administrators, and others interested in using bibliometrics or assessing the relevance of bibliometric results.) | Website | N/A | No | No | No | No | 3 | No |
| Cabezas-Clavijo et al.<br>(2021) | Spain | The main goal of this study is to outline a series of recommendations for bibliometricians, consultants and research support librarians when drafting bibliometric reports in their institutions. | Guidance Paper | Research Methods and Assessment | No | No | No | No | 15 | Yes |
| Cabezas-Clavijo et al. (2022) | Spain | Using Sustainability (the journal outside the IS area that publishes the most bibliometric articles) as a case study, this study uses content analysis to determine various parameters relating to the methodological rigour and reproducibility of the papers published in this journal in 2019 and 2020. | Conference Guidance Paper | N/A | No | No | Partially | No | 3 | No |
| Cabezas-Clavijo et al. (2023) | Spain | The aim of this article is to explore up to seven parameters related to the methodological quality and reproducibility of thematic bibliometric research published in the two most productive journals in bibliometrics, Sustainability (a journal outside the discipline) and Scientometrics, the flagship journal in the field. | Research Paper | Data and Information Science | No | No | Partially | No | 7 | Yes |
| Dey (2023) | India | Not explicitly provided in the main body of text.<br>(Title: Guidelines for Crafting PURPOSEFUL Bibliometric Reviews for SAJBMC) | LinkedIn | N/A | No | No | No | No | 5 | Yes |
| Donthu et al. (2021) | USA | This paper aims to offer (1) an overview of the bibliometric methodology and (2) step-by-step guidelines for conducting bibliometric analysis for business research. | Guidance Paper | Business | No | No | No | Yes | 5 | Yes |
| Gan et al. (2022) | China | The aim of this paper is to summarize bibliometric and mapping knowledge domain (MKD) methodologies, databases that should be used, useful software, their limitations and potential application to traditional, complementary, and integrative medicine (TCIM) research. | Guidance Paper | Complementary and Integrative Health Care | Yes<br>Mapping knowledge domains was extensively discussed in addition to bibliometric analysis | No | No | No | 2 | Yes |
| Glänzel & Schoepflin (1994) | Hungary | The authors plead for integrative and interdisciplinary research approaches, for reinforcing fundamental, methodological and experimental research programs in scientometrics, for independent funding of research, and for an enhancement of scientometric databases. The need for acknowledged technical and scientific standards in research and publication is stressed. Finally, the establishment of a Code of Etlu'cs for the field of scientometrics is proposed. | Guidance Paper | Scientific and Technological Information, Scientometrics | No | No | No | No | 4 | Yes |
| Glänzel (1996) | Hungary | The need for standardisation in bibliometric research and technology is discussed in the context of failing communication within the scientific community, the unsatisfactory impact of bibliometric research outside the community and the observed incompatibility of bibliometric indicators produced by different institutes. | Guidance Paper | Scientific and Technological Information, Scientometrics | No | No | No | No | 6 | No |
| Hicks et al. (2015) | USA | We offer this distillation of best practice in metrics-based research assessment so that researchers can hold evaluators to account, and evaluators can hold their indicators to account. We therefore present the Leiden Manifesto. | Guidance Paper | Interdisciplinary | No | Yes | No | No | 6 | No |
| Koo et al. (2023) | Canada, Taiwan | The present study aimed to investigate the reporting practices of bibliometric research related to health and medicine based on a guideline "Preferred Reporting Items for Bibliometric Analysis (PRIBA)" proposed in this study. | Reporting Guideline | Interdisciplinary | No | No | Partially | No | 15 | No |
| Lim & Kumar (2023) | Malaysia | This article proposes a sensemaking approach that transitions researchers from mere description to proactive interpretation of bibliometric results, transforming raw information into informed insights. | Guidance Paper | Business | Yes<br>Sense-making as a theoretical lens | Yes | No | No | 6 | Yes |
| Linnenluecke et al. (2020) | Australia | In this article, we detail methodological steps for how researchers can conduct systematic literature reviews and offer examples of bibliometric approaches to visualise results. | Guidance Paper | Management, Accounting, Finance, Social Sciences | Yes<br>Systematic Reviews | No | No | Yes | 1 | Yes |
| Montazeri et al. (2023) | Iran | This study aimed to provide a guideline for reporting bibliometric reviews [of biomedical literature] | Reporting Guideline | Systematic Reviews | No | No | Yes | No | 14 | No |
| Mukherjee et al. (2022) | USA | [We address] the following research question: How can bibliometric research contribute to theory and practice? | Editorial | Management, Business | No | No | No | No | 9 | Yes |
| Pendlebury (2010) | USA | Not explicitly provided.<br>(Thomson Reuters offers some practical advice on how to approach and evaluate research performance using quantitative indicators, such as those we and others make available [through a ten-item checklist]) | White Paper | N/A | No | Yes | No | No | 4 | Yes |
| Romanelli et al. (2021) | Brazil | First, to discuss four big challenges researchers may face when conducting bibliometric studies and how to deal with them. We go forward discussing some primary questions researchers may address with bibliometric mapping, drawing examples from four authorial case studies. Second, we discuss how bibliometric studies can improve in quality by suggesting best practices based on principles of systematic reviews and maps. | Guidance Paper | Environmental Sciences | No | No | No | Yes | 6 | Yes |
| Technical University of Munich (2023) | Germany | The San Francisco Declaration on Research Assessment (DORA) [...] contains recommendations for a fair and transparent use of bibliometrics with regard to the assessment of research performance. | University Declaration | N/A | No | Yes | No | No | 8 | Yes |
| University of Oulu (Last Updated 2024) | Finland | Not explicitly provided by source.<br>Provides recommendations on how to use publication metrics responsibly | Website | N/A | No | No | No | No | 3 | No |
| Webster (2017) | USA | The purpose of this paper is to summarize and provide context to the recently published Leiden Manifesto, a document written by leading bibliometric researchers, which proposes ten principles that should guide the use of bibliometric tools and indicators in research evaluation. | Opinion Piece | Performance Measurement and Assessment | No | Yes | No | No | 5 | Yes |
| Zupic et al. (2015) | Slovenia | The purpose of this article is to develop a meaningful single-source reference for management and organization of scholars interested in bibliometric methods. | Guidance Paper | Organizational Sciences | No | No | Partially | No | 5 | No |

<sup>a</sup> Evidence-based recommendations were measured by the sources's adherence to EQUATOR's "How to develop a reporting guideline" toolkit (EQUATOR, 2018c); 'Yes' indicates that the document conducted a systematic review of the literature and/or a consensus exercise, 'Partially' indicates that the document conducted a review of at least one bibliometric database and/or journal, 'No' indicates that the document was opinion-based.

The first authors of the included documents originated from a variety of countries including the USA (n = 6), Spain (n = 3), Germany (n = 2), Hungary (n = 2), as well as Australia, Brazil, Canada, China, Finland, India, Iran, Malaysia, and Slovenia (all n = 1). One author was affiliated with institutions in both Taiwan and Canada. Documents were also published in a variety of journals with the following scopes of interest: business (n = 2), interdisciplinary (n = 2), management (n = 2), research/performance methods and assessment (n = 2), scientometrics (n = 2), complementary and integrative healthcare (n = 1), data and information science (n = 1), environmental sciences (n = 1), natural sciences (n = 1), organizational sciences (n = 1), and systematic reviews (n = 1). Seven sources were not published in a journal and were consequently not assigned a journal subject scope.

Of the 23 included sources, 14 provided a conceptual framework to understand bibliometric analyses and five sources conducted a bibliometric analysis as part of their document to provide reporting recommendation examples. Four sources had a special focus on the use of bibliometrics for the hiring, promotion, and tenure of researchers at institutions, and three sources had a special focus on non-bibliometric subject matter (i.e., mapping knowledge domains [n = 1], sensemaking as a theoretical lens [n = 1], and systematic reviews [n = 1]).

In terms of the evidence-based nature of bibliometric reporting recommendations, sources were categorised based on their adherence to EQUATOR’s “How to develop a reporting guideline” toolkit (EQUATOR, 2018c). This toolkit suggests that researchers creating a reporting recommendation guideline should conduct a ‘thorough literature review’ (EQUATOR, 2018b) along with a ‘face-to-face consensus meeting’ (EQUATOR, 2018a). Seventeen out of the 23 included documents were opinion-based and did not adhere to EQUATOR’s toolkit when creating bibliometric reporting recommendations (i.e., they did not show evidence of a systematic review of the literature and/or conduct a consensus exercise). Two sources (Cabezas-Clavijo et al., 2023; Cabezas-Clavijo et al., 2022) specified some degree of literature review to inform their recommendations, but these searches were limited to a few journals. For instance, Cabezas-Clavijo et al. (2022) evaluated 204 bibliometric analysis studies specifically from the journal *Sustainability* (Sustainability, n.d.). Three documents (Boyack et al., 2022; Koo & Lin, 2023; Zupic & Čater, 2015) demonstrated evidence of conducting searches in at least one bibliographic database but did not describe any consensus process. Only one (Montazeri et al., 2023) adhered to EQUATOR’s toolkit by conducting a systematic search of the literature through bibliographic databases and conducting a Delphi consensus meeting to create reporting recommendations in the form of a preliminary Guideline for Reporting Bibliometric Reviews of the Biomedical Literature (BIBLIO).

### 3.3. Data Extraction and Item Synthesis

Bibliometric reporting recommendations extracted from the included sources were synthesised to create a preliminary GLOBAL reporting guideline with 32 candidate items (see **Table 3** for the list). In the guideline, items were sub-categorised according to the section that the bibliometric reporting recommendation pertained to: ‘title’ (1 item), ‘abstract’ (1 item), ‘introduction’ (5 items), ‘methods’ (13 items), ‘results’ (4 items), ‘discussion’ (5 items), and ‘other’ (3 items). The included documents made between one and 15 bibliometric reporting recommendations each (see **Table 2** for exact values). The complete data extraction form and the data in support of the GLOBAL candidate items from included articles can be found at Ng et al. (2024).

**Table 3:** Preliminary GLOBAL Recommendation List.

| Reporting Items |  | Studies that Provide Guidance |  |
| --- | --- | --- | --- |
| Title |  | Number | References |
| 1.1 | In the title, identify the study as a bibliometric analysis and indicate the time period and key issues/topic. | 2 | (Koo & Lin, 2023; Montazeri et al., 2023) |
| Abstract |  |  |  |
| 2.1 | Abstract should be reflective of the bibliometric analysis, including scope, data collection, analysis, and results. | 2 | (Koo & Lin, 2023; Montazeri et al., 2023) |
| Introduction |  |  |  |
| 3.1 | Situate the bibliometric analysis within the context of relevant pre-existing literature, identifying the gap in literature. | 4 | (Boyack et al., 2022; Glänzel & Schoepflin, 1994; Montazeri et al., 2023; Mukherjee et al., 2022) |
| 3.2 | Define the aim, scope, rationale, and/or objective of the bibliometric analysis. | 7 | (Byl et al., 2016; Cabezas-Clavijo & Torres-Salinas, 2021; Dey, 2023; Donthu et al., 2021; Hicks et al., 2015; Koo & Lin, 2023; Montazeri et al., 2023) |
| 3.3 | Define the research question. | 3 | (Byl et al., 2016; Dey, 2023; Zupic & Čater, 2015) |
| 3.4 | Clearly define all relevant terms and definitions used within the bibliometric analysis. | 1 | (Glänzel, 1996) |
| 3.5 | Describe the intended target audience of the bibliometric analysis (e.g., researchers, public, media, etc.). Describe the ways in which the information included in the report may be used for the target audience. | 3 | (Boyack et al., 2022; Cabezas-Clavijo & Torres-Salinas, 2021; Mukherjee et al., 2022) |
| Methods |  |  |  |
| 4.1 | Describe the bibliometric methods used. | 8 | (Cabezas-Clavijo & Torres-Salinas, 2021; Glänzel, 1996; Lim & Kumar, 2023; Montazeri et al., 2023; Mukherjee et al., 2022; Romanelli et al., 2021; Technical University of Munich, 2023; Zupic & Čater, 2015) |
| 4.2 | Define the units of analysis that are analysed (i.e., micro-, meso-, and macro-level) in the bibliometric analysis (e.g., countries, institutions, authors). | 1 | (Cabezas-Clavijo & Torres-Salinas, 2021) |
| 4.3 | Describe the bibliometric data collection methods, including any limitations. | 4 | (Cabezas-Clavijo & Torres-Salinas, 2021; Hicks et al., 2015; Romanelli et al., 2021; Webster, 2017) |
| 4.4 | Describe the databases and data sources used, including any limitations. | 11 | (Bornmann et al., 2014; Byl et al., 2016; Cabezas-Clavijo et al., 2023; Cabezas-Clavijo et al., 2022; Cabezas-Clavijo & Torres-Salinas, 2021; Glänzel, 1996; |
|  |  |  | Hicks et al., 2015; Koo & Lin, 2023; Montazeri et al., 2023; Romanelli et al., 2021; Technical University of Munich, 2023) |
| 4.5 | Present the full search strategies for all databases used, including any filters and limits that were applied. | 13 | (Bornmann et al., 2014; Cabezas-Clavijo et al., 2023; Cabezas-Clavijo et al., 2022; Cabezas-Clavijo & Torres-Salinas, 2021; Dey, 2023; Gan et al., 2022; Glänzel, 1996; Koo & Lin, 2023; Montazeri et al., 2023; Pendlebury, 2010; Romanelli et al., 2021; Technical University of Munich, 2023; Zupic & Čater, 2015) |
| 4.6 | Describe the data collection time frame. | 3 | (Bornmann et al., 2014; Cabezas-Clavijo et al., 2023; Technical University of Munich, 2023) |
| 4.7 | Describe the search results and selection processes (e.g., inclusion/exclusion). If applicable, use a flow diagram. | 2 | (Koo & Lin, 2023; Montazeri et al., 2023) |
| 4.8 | Describe the data cleaning methods, including any limitations. | 0 | - |
| 4.9 | Describe the bibliometric data analysis methods used. | 2 | (Cabezas-Clavijo et al., 2023; Montazeri et al., 2023) |
| 4.10 | Specify the analytical software used and the parameter settings selected. | 2 | (Gan et al., 2022; Koo & Lin, 2023) |
| 4.11 | Describe the bibliometric indicators used. | 8 | (Cabezas-Clavijo et al., 2023; Cabezas-Clavijo & Torres-Salinas, 2021; Glänzel, 1996; Koo & Lin, 2023; Pendlebury, 2010; Technical University of Munich, 2023; University of Oulu, n.d.; Webster, 2017) |
| 4.12 | If applicable, define the calculations/formulas used for indicators in the bibliometric analysis. | 4 | (Cabezas-Clavijo et al., 2023; Cabezas-Clavijo & Torres-Salinas, 2021; Technical University of Munich, 2023; Webster, 2017) |
| 4.13 | Provide sufficient detail in the bibliometric analysis manuscript to ensure full replicability/transparency of methods. | 14 | (Boyack et al., 2022; Cabezas-Clavijo et al., 2023; Cabezas-Clavijo et al., 2022; Cabezas-Clavijo & Torres-Salinas, 2021; Glänzel, 1996; Glänzel & Schoepflin, 1994; Hicks et al., 2015; Lim & Kumar, 2023; Mukherjee et al., 2022; Pendlebury, 2010; Romanelli et al., 2021; Technical University of Munich, 2023; University of Oulu, n.d.; Webster, 2017) |
| **Results** |  |  |  |
| 5.1 | Describe the results and key findings. | 4 | (Boyack et al., 2022; Lim & Kumar, 2023; Webster, 2017; Zupic & Čater, 2015) |
| 5.2 | Describe the results of bibliometric analysis techniques used. | 3 | (Hicks et al., 2015; Koo & Lin, 2023; Montazeri et al., 2023) |
| 5.3 | Visualize the results through the use of figures, graphs, and/or tables. Ensure the visualizations are simple and easy to interpret. Aesthetic bibliometric visualization should not replace a rigorous bibliometric analysis. | 7 | (Boyack et al., 2022; Cabezas-Clavijo & Torres-Salinas, 2021; Dey, 2023; Donthu et al., 2021; Koo & Lin, 2023; Linnenluecke et al., 2020; Montazeri et al., 2023) |
| 5.4 | If applicable, report the uncertainty/dispersion/heterogeneity depending on the type of analysis and error values of bibliometric indicators. | 1 | (Hicks et al., 2015) |
| **Discussion** |  |  |  |
| 6.1 | Summarize and discuss study findings. | 8 | (Bornmann et al., 2014; Boyack et al., 2022; Donthu et al., 2021; Koo & Lin, 2023; Lim & Kumar, 2023; Montazeri et al., 2023; Mukherjee et al., 2022; Zupic & Čater, 2015) |
| 6.2 | Elaborate on the applicability and implications of study findings. | 8 | (Boyack et al., 2022; Cabezas-Clavijo & Torres-Salinas, 2021; Dey, 2023; Donthu et al., 2021; Lim & Kumar, 2023; Montazeri et al., 2023; Mukherjee et al., 2022; Pendlebury, 2010) |
| 6.3 | Provide context for the results of the bibliometric analysis and situate the study findings in existing literature. | 6 | (Cabezas-Clavijo & Torres-Salinas, 2021; Donthu et al., 2021; Glänzel & Schoepflin, 1994; Koo & Lin, 2023; Mukherjee et al., 2022; Zupic & Čater, 2015) |
| 6.4 | Discuss the strengths, limitations, and potential biases of the bibliometric analysis. | 7 | (Cabezas-Clavijo & Torres-Salinas, 2021; Koo & Lin, 2023; Montazeri et al., 2023; Mukherjee et al., 2022; Romanelli et al., 2021; Technical University of Munich, 2023; University of Oulu, n.d.) |
| 6.5 | Identify future directions for research. | 1 | (Mukherjee et al., 2022) |
| **Other** |  |  |  |
| 7.1 | Disclose any existing or potential conflicts of interest and/or sources of financial or non-financial support. | 3 | (Boyack et al., 2022; Cabezas-Clavijo & Torres-Salinas, 2021; Koo & Lin, 2023) |
| 7.2 | Describe the availability and accessibility of data. | 1 | (Koo & Lin, 2023) |
| 7.3 | Use references and citations to support statements and methods used. | 1 | (Lim & Kumar, 2023) |

The most frequently addressed item (14 times) was making sure that methods are reported in a replicable and transparent manner (**Table 3**). Ensuring that full database search strategies are presented in bibliometric analyses, including any filters and limitations applied, was addressed in 13 documents. Further, describing the databases and data sources used, with limitations, was cited in 11 sources. Alternatively, six items on the preliminary GLOBAL recommendation list were only mentioned by one study each: 1) defining all relevant terms and definitions used in the bibliometric analysis; 2) defining the units of analysis; 3) reporting the uncertainty/dispersion/heterogeneity and error values of bibliometric indicators when applicable; 4) identifying future directions for research; 5) describing the availability and accessibility of the data; and 6) using references and citations to support statements and methods used. One item (‘describe the data cleaning methods, including any limitations’) was not described in any literature but was added to the preliminary reporting list based on the opinions of bibliometric experts who are part of the research team.

## 4. Discussion

The objective of this scoping review was to identify recommendations for the reporting of bibliometric analyses from journals, preprint servers, and grey literature to inform the development of the GLOBAL, a reporting guideline for bibliometric analyses. Of the 48,750 records screened, a total of 23 documents met the inclusion criteria (see **Figure 1** for PRISMA flow chart and **Table 2** for included study characteristics). A 32-item preliminary GLOBAL recommendation list (**Table 3**) was produced through this analysis. One of the 32 items was added as the result of expert opinion from the internal GLOBAL steering committee, while the remaining 31 items were created based on recommendations in the literature.

Currently, limited robust and universal reporting recommendations exist for bibliometric analyses. Results from the present review further support this idea. Seventeen of the 23 included sources were based on opinion as opposed to rigorous methodologies (e.g., did not report conducting a review of the literature to inform recommendations or conducting a face-to-face consensus process), with only two published in scientometric journals. Two of the included documents completed a systematic review of the literature in at least one bibliographic database, while three of the included documents conducted a partial review of the literature in one or more journals. Only one included source adhered to EQUATOR’s recommendations (EQUATOR, 2018c) by conducting a systematic literature review and a Delphi consensus process to create a reporting guideline for bibliometric analyses, referred to as BIBLIO (Montazeri et al., 2023). However, this reporting guideline has some limitations. For instance, only eleven experts were involved in the consensus project, which may not adequately reflect the opinions of the broader international bibliometrics community. Also, some details were missing from the reporting of the study, which makes assessing its validity and reliability difficult (e.g., limited information was provided about how the literature review was conducted, and descriptive statistics of the results for the individual Delphi voting rounds were not reported). The overarching paucity of robust and transparent research present within this area consequently supports the need for the GLOBAL.

When developing the GLOBAL, the frequency with which different items were addressed in bibliometric studies varied, highlighting key areas of emphasis and their perceived importance within the research community. Transparency in reporting methods, as indicated in the 14 included documents, emerged as the most frequently mentioned item. The importance of research transparency has similarly been highlighted in the literature.

Transparency enhances the credibility of scientific papers through replication and verification of results, which in turn, increases the robustness of scientific knowledge (Hardwicke et al., 2022; Malički et al., 2023; Serghiou et al., 2021). In contrast, despite its crucial role in confirming the quality and validity of an analysis (Guo et al., 2023; Ridzuan & Wan Zainon, 2019), data cleaning methods (i.e., removing duplicates or fixing incomplete datasets to ensure that the data gathered accurately reflects the situation being studied) were not mentioned in any of the reviewed sources. This discrepancy suggests a potential oversight or underappreciation of this aspect within current bibliometric reporting practices, reinforcing the need for additional bibliometric reporting resources, such as the GLOBAL.

The resulting GLOBAL reporting guideline is ultimately intended to aid bibliometricians, librarians, policymakers, research evaluators, and researchers in the assessment and use of bibliometric analyses, as well as authors, editors, and peer reviewers who are conducting or reviewing bibliometric analyses. By creating this guideline, we aim to facilitate an increase in the quality of reporting of bibliometric analyses (Moher et al., 2010) and ensure the completeness and transparency of their published analyses (Gagnier et al., 2013). As bibliometric analyses become increasingly popular (Donthu et al., 2021; Ellegaard & Wallin, 2015) and bibliometric databases more accessible (Barcelona Declaration on Open Research Information, 2023), it is important to establish standard reporting guidelines for these types of studies to increase the reliability and accuracy of published bibliometric research.

In subsequent steps, the preliminary 32-item GLOBAL list (see **Table 3**) will be further developed through a Delphi study that is currently underway. Relevant stakeholders were invited to participate in an online survey, which will be followed by an international consensus meeting taking place in Berlin in September 2024 following the 28th International Conference on Science, Technology and Innovation Indicators. Once completed, the GLOBAL will serve as the first guideline developed for the reporting of bibliometric analyses through international, multi-stakeholder consensus. In accordance with the approach followed by the Consolidated Standards of Reporting Trials (CONSORT) statement (Altman, 1996), we intend for this guideline to be clear and easy to follow, such that, ‘[r]eaders should not have to infer what was probably done; they should be told explicitly.’ While the GLOBAL checklist is intended to provide clear recommendations to increase the reproducibility of bibliometric analyses, we acknowledge that practical considerations (e.g., journal requirements or concision) may hinder the ability for researchers to provide the full scope of information needed to meet ideal standards of reproducibility and transparency.

Nevertheless, we anticipate that the widespread adoption of the GLOBAL will result in more thorough, accurate, and transparent reporting of bibliometric studies.

### 4.1. Strengths and Limitations

The title/abstract and full-text screening of the records, as well as the extraction of recommendations from them, were conducted independently and in duplicate, serving as a strength of the study. Furthermore, we screened a wide range of literature, including journals, preprint servers, and grey literature sources, to ensure a thorough review of the recommendations on reporting bibliometric studies. However, the inclusion of records only available in English serves as a study limitation, as non-English language documents with potential reporting recommendations may have been overlooked. Furthermore, while we explored both publicly available search results and those accessible through our university library systems and interlibrary loans, we acknowledge that there still exists a possibility that not all relevant recommendations for reporting bibliometric analysis were captured.

## 5. Conclusion

The aim of this scoping review is to identify bibliometric reporting recommendations from various forms of literature to develop an initial list of candidate items for the GLOBAL reporting guideline. Ultimately, 23 records met the eligibility criteria out of the 48,750 documents that were reviewed. With most recommendations being opinion-based (n = 17), there is a lack of robust standard reporting guidelines for bibliometric analyses. A 32-item preliminary GLOBAL recommendation list was created through this scoping review process and will be further developed in a Delphi study. Once finalised, the GLOBAL will function as a standardised guideline for reporting bibliometric analyses and, if widely adopted, is expected to result in higher-quality reporting of bibliometric studies.

## List of Abbreviations

ACCORD: ACcurate COnsensus Reporting Document
BIBLIO: Guideline for Reporting Bibliometric Reviews of the Biomedical Literature
CONSORT: Consolidated Standards of Reporting Trials
DAISY: DistillerSR Artificial Intelligence System
DOI: Digital Object Identifier
EQUATOR: Enhancing the QUAlity and Transparency Of health Research
GLOBAL: Guidance List for the repOrting of Bibliometric AnaLyses
JBI: Joanna Briggs InstituteOSF: Open Science Framework
PCC: Participants, Concept, Context
PRESS: Peer Review of Electronic Search Strategies
PRISMA: Preferred Reporting Items for Systematic Reviews and Meta-Analyses
PRISMA-P: Preferred Reporting Items for Systematic Review and Meta-Analysis Protocols
PRISMA-ScR: Preferred Reporting Items for Systematic Reviews and Meta-Analyses extension for Scoping Reviews
SCI-EXPANDED: Science Citation Index Expanded SSCI: Social Sciences Citation Index
URL: Uniform Resource Locator

## Declarations

### Ethics Approval and Consent to Participate

This study involved a systematic review of peer-reviewed literature only; it did not require ethics approval or consent to participate.

### Consent for Publication

All authors consent to this manuscript’s publication.

### Availability of Data and Materials

All relevant data are included in this manuscript or posted on the Open Science Framework: https://doi.org/10.17605/OSF.IO/WYP63 (Ng et al., 2024).

### Authors’ Contributions

JYN: Conceptualization, Data curation, Formal Analysis, Funding acquisition, Investigation, Methodology, Writing – original draft

HL: Data curation, Formal Analysis, Investigation, Methodology, Writing – original draft MM: Data curation, Formal Analysis, Investigation, Methodology, Writing – original draft NS: Data curation, Formal Analysis, Investigation, Methodology, Writing – original draft DS: Formal Analysis, Investigation, Writing – review & editing

APA: Formal Analysis, Investigation, Writing – review & editing

MSabé: Conceptualization, Formal Analysis, Investigation, Methodology, Writing – review & editing

MSolmi: Conceptualization, Formal Analysis, Investigation, Methodology, Writing – review & editing

LW: Conceptualization, Formal Analysis, Investigation, Methodology, Writing – review & editing

SH: Conceptualization, Formal Analysis, Investigation, Methodology, Writing – review & editing

DM: Conceptualization, Formal Analysis, Investigation, Methodology, Writing – review & editing

## Data Availability

All relevant data are included in this manuscript or posted on the Open Science Framework.

https://doi.org/10.17605/OSF.IO/WYP63

## Acknowledgements

We gratefully acknowledge Chaomei Chen and Sanam Ebrahimzadeh for their contributions to the study design and protocol development. We also gratefully acknowledge Kaitlin Fuller for her contributions in PRESSing the bibliographic database search strategies developed by APA.

## Appendices

### Appendix A: Search Strategies for Bibliometric Analysis Recommendations

All searches were retrieved July 28, 2023.

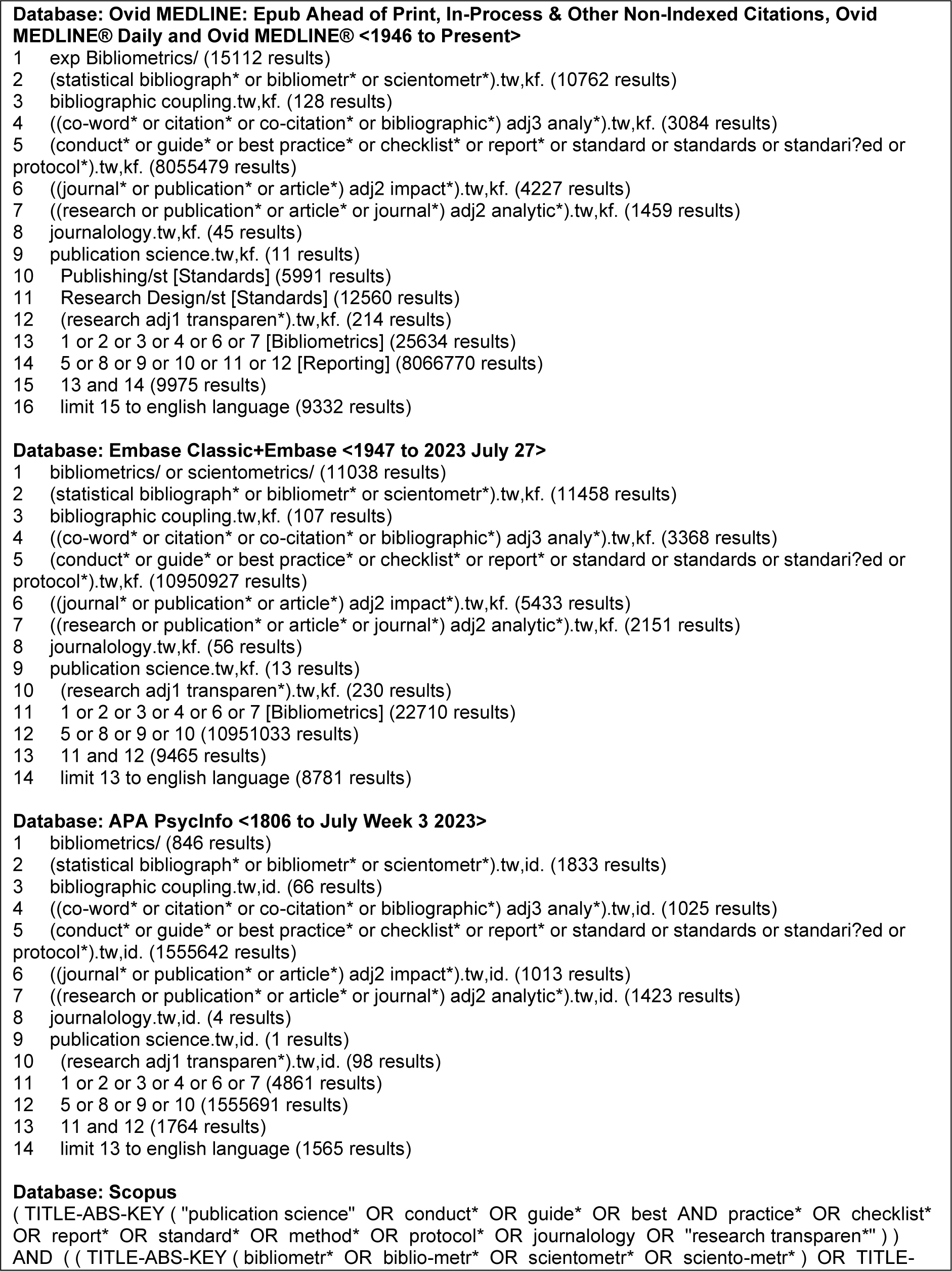
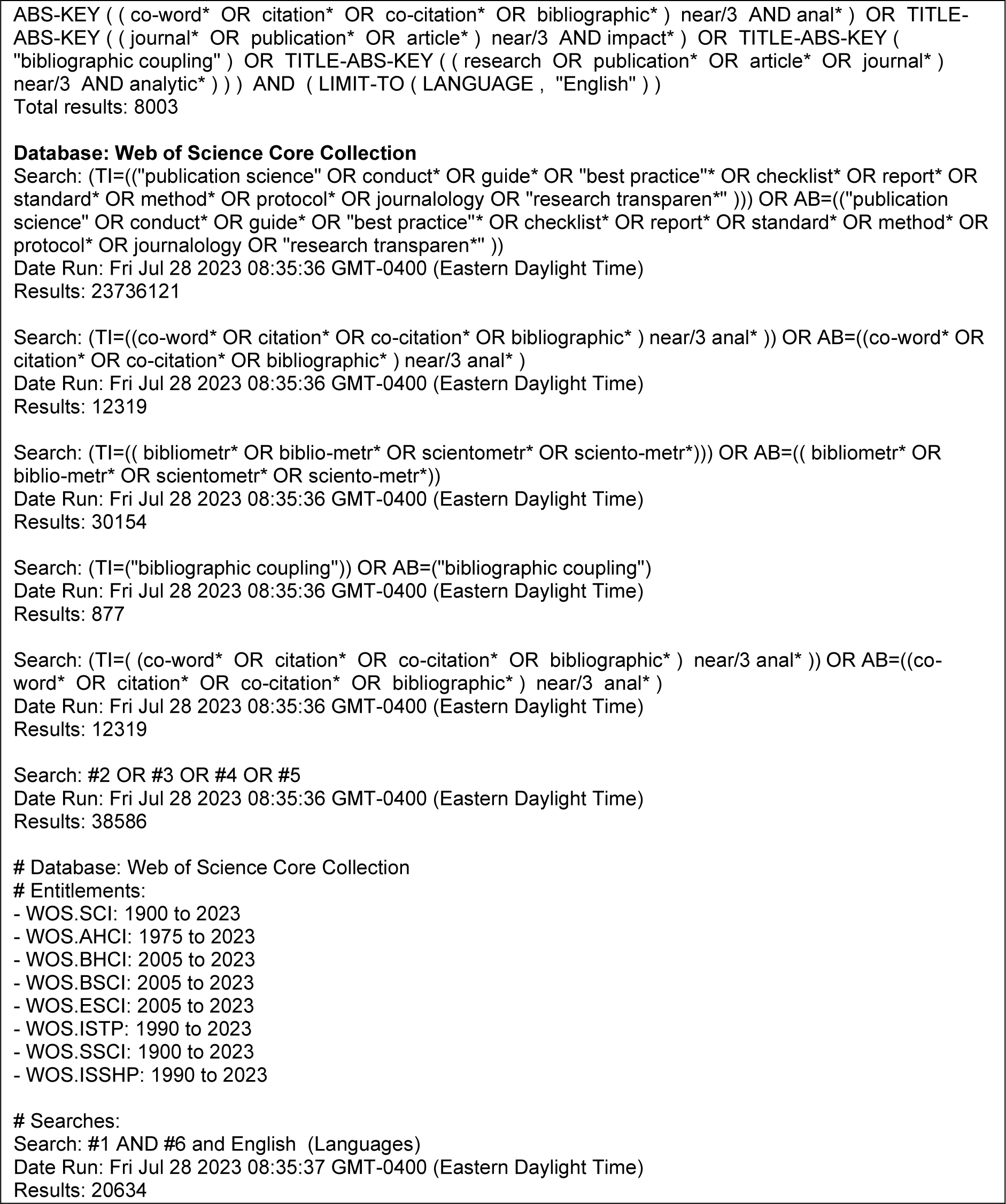

### Appendix B: Grey Literature Sources for Bibliometric Analysis Recommendations

Recommendations
| Source | Website |
| --- | --- |
| International Society for Scientometrics and Informetrics (ISSI) | https://www.issi-society.org/ |
| International Network of Research Management Societies (INORMS) | https://inorms.net/ |
| Directorate for Science, Technology and Innovation (STI) | https://www.oecd.org/sti/ |
| European Network of Indicator Designers (ENID) | https://enid-europe.eu/ |
| Association for Information Science and Technology (ASIS&T) | https://www.asist.org/ |
| International Association for Social Science Information Service and Technology (IASSIST) | https://iassistdata.org/ |
| Centre for Science and Technology Studies (CWTS) | https://www.cwts.nl/ |
| Science-Metrix | https://www.science-metrix.com/ |
| Google Searches<br>("guidance on reporting bibliometric analyses" and searching the top 100 hits) | https://www.google.ca/ |

